# Mitochondria-targeted antioxidant supplementation augments acute exercise-induced increases in muscle PGC1α mRNA and improves training-induced increases in peak power independent of mitochondrial content and function in untrained middle-aged men

**DOI:** 10.1101/2021.12.05.21267323

**Authors:** S. C. Broome, T. Pham, A. J. Braakhuis, R. Narang, H. W. Wang, A. J. R. Hickey, C. J. Mitchell, T. L. Merry

**Author notes:** <u>Corresponding author:</u> Sophie Broome, Department of Nutrition, School of Medical Sciences, The University of Auckland, Private Bag 92019, Auckland, New Zealand.

## Abstract

The role of mitochondrial ROS production and signalling in muscle adaptations to exercise training has not been explored in detail. Here we investigated the effect of supplementation with the mitochondria-targeted antioxidant MitoQ on a) the skeletal muscle mitochondrial and antioxidant gene transcriptional response to acute high-intensity exercise and b) skeletal muscle mitochondrial content and function following exercise training. In a randomised, double-blind, placebo-controlled, parallel design study, 23 untrained men (age: 44 ± 7 years, VO_2peak_: 39.6 ± 7.9 ml/kg/min) were randomised to receive either MitoQ (20 mg/d) or a placebo for 10 days before completing a bout of high-intensity interval exercise (cycle ergometer, 10 × 60 s at VO_2peak_ workload with 75 s rest). Blood samples and vastus lateralis muscle biopsies were collected before exercise and immediately and 3 hours after exercise. Participants then completed high-intensity interval training (HIIT; 3 sessions per week for 3 weeks) and another blood sample and muscle biopsy were collected. MitoQ supplementation augmented acute exercise-induced increases in skeletal muscle mRNA expression of the major regulator of proteins involved in mitochondrial biogenesis peroxisome proliferator-activated receptor gamma coactivator 1-alpha (PGC1-α). Despite this, training-induced increases in skeletal muscle mitochondrial content were unaffected by MitoQ supplementation. HIIT-induced increases in VO_2peak_ and 20 km time trial performance were also unaffected by MitoQ while MitoQ augmented training-induced increases in peak power achieved during the VO_2peak_ test. These data suggest that MitoQ supplementation enhances the effect of training on peak power, which may be related to the augmentation of skeletal muscle PGC1α expression following acute exercise. However, this effect does not appear to be related to an effect of MitoQ supplementation on HIIT-induced mitochondrial biogenesis in skeletal muscle and may therefore be the result of other adaptations mediated by PGC1α.

## INTRODUCTION

Exercise-induced stressors trigger an acute and transient upregulation of gene expression in skeletal muscle that, if reinforced by repeated exercise bouts as part of an exercise training program, results in phenotypic adaptations that improve skeletal muscle function [1, 2]. In recent years, reactive oxygen species (ROS) have become recognised as important exercise-stress signalling intermediates that regulate the adaptive response to exercise training [3-5]. Since high ROS levels can impair muscle function [6, 7], and are therefore thought to limit exercise performance, antioxidant supplements are commonly consumed by athletes in an effort to limit exercise-induced increases in oxidative stress [8]. While antioxidant supplementation may have benefits for acute performance in some instances [9, 10], generalised antioxidant supplementation has been shown to impair transcriptional responses to acute exercise [4, 5, 11-13], resulting in attenuated training-induced performance [11] and metabolic [5] benefits.

A central adaptation to regular endurance exercise training is increased skeletal muscle mitochondrial volume and enhanced antioxidant capacity [14, 15]. The redox sensitive transcription factor peroxisome proliferator-activated receptor gamma coactivator 1-alpha (PGC1α) increases with exercise, is involved in coordinating mitochondrial biogenesis, and upregulates expression of antioxidant enzymes, including superoxide dismutase (SOD), glutathione peroxidase (GPx), and catalase [16]. Studies in mice and humans have shown that supplementation with the general antioxidants vitamin C and E attenuates exercise induced-increases in PGC1α [5, 11, 12], which may explain why some studies report that general antioxidant supplementation negates some of the beneficial effects of exercise, such as increases in skeletal muscle mitochondrial content and antioxidant capacity [5, 11, 12], and improvements in insulin sensitivity [5].

While the major sources of ROS during exercise appear to be non-mitochondrial (i.e. NADPH oxidase (NOX) enzymes [17]), there is also evidence of changes in the mitochondrial redox environment during exercise with reports of increased skeletal muscle mitochondrial DNA (mtDNA) damage [18] and oxidation of the mitochondrial matrix localised peroxiredoxin (Prx) 3 [19]. To date, most human studies investigating the effect of antioxidant supplementation on skeletal muscle responses to exercise have used general antioxidants, such as N-acetylcysteine or a combination of vitamin C and E [5, 11, 12, 20], which may result in indiscriminate neutralisation of ROS from multiple intracellular sources. Therefore, the role of mitochondrial ROS production and signalling during exercise in muscle adaptations to exercise training is yet to be explored in detail.

Supplementation with mitochondria-targeted coenzyme Q10 (mitoquinone; MitoQ) has previously been shown to attenuate exercise-induced increases in mtDNA damage in human skeletal muscle [18]. MitoQ consists of a ubiquinone moiety, which prevents lipid peroxidation by acting as a chain-breaking antioxidant and recycling the α-tocopheroxyl radical to its active form [21, 22], conjugated to a triphenylphosphonium cation to facilitate accumulation within mitochondria [23, 24]. Here we investigated the effect of MitoQ supplementation on a) skeletal muscle mitochondrial and antioxidant gene transcriptional response to acute high-intensity exercise and b) skeletal muscle mitochondrial content and function following exercise training.

## MATERIALS AND METHODS

### Participants

Twenty-five healthy middle-aged men were recruited via advertisements and provided written informed consent to participate in the trial (Supplementary figure 1). A middle-aged cohort was studied as use of dietary supplements has been shown to increase with age [25]. Two participants withdrew from the study before the acute exercise trial due to time commitments and one participant withdrew from the study after completion of the acute exercise trial due to illness. Results are presented for the 23 participants who completed the acute exercise trial. Participant characteristics are given in Table 1. All participants reported as non-smokers, free of injury and chronic illnesses including cardiovascular and metabolic disease, free from antioxidant supplementation for > 6 weeks before enrollment in the study and completed < 2 h physical activity/week. The study was approved by the Northern Health and Disability Ethics Committee (New Zealand), registered with the Australian New Zealand Clinical Trial Registry (ACTRN12618000238279) on 14^th^ February 2018 and conducted in accordance with the ethical standards laid down in the Declaration of Helsinki.

**Table 1.** Participant characteristics.

| Characteristics | Placebo group (n=11) | MitoQ group (n=12) | p value |
| --- | --- | --- | --- |
| Age (years) | 44 ± 8 | 44 ± 7 | 0.972 |
| Height (cm) | 175.9 ± 0.1 | 178.0 ± .01 | 0.525 |
| Weight (kg) | 80.4 ± 16.7 | 83.3 ± 13.8 | 0.667 |
| BMI (kg/m<sup>2</sup>) | 25.7 ± 3.3 | 26.1 ± 2.9 | 0.754 |
| V̇O<sub>2peak</sub> (ml/kg/min) | 39.1 ± 7.4 | 37.0 ± 7.9 | 0.531 |
| Peak power at VO<sub>2peak</sub> (W) | 237 ± 46 | 232 ± 34 | 0.825 |

Values are mean ± SD

### Exercise study protocol

The study was a double-blind, placebo-controlled parallel design (Figure 1a). Participants completed a ramp-incremental (starting at 60 W and increasing by 15 W per minute) exercise test to voluntary exhaustion using an electromagnetically braked cycle ergometer (Velotron, RacerMate, Seattle, WA) with open-circuit spirometry (Parvo Medics True One 2400, Sandy, UT) to determine peak oxygen uptake (VO_2peak_) and peak power at VO_2peak_. Participants were familiarised to the time trial, which involved cycling 20 km in the shortest time possible, before returning at least 72 hours later to complete the time trial. The time trial mode of the cycle ergometer allowed for the use of self-selected gearing and participants received visual feedback on distance completed, but not time elapsed. Participants then supplemented with MitoQ (20 mg/day) or a placebo for 10 days before completing the acute exercise trial followed by nine sessions of high-intensity interval training (HIIT). On the day of the acute exercise trial, participants arrived at the laboratory following a 10 hour overnight fast and completed 10 × 60 second cycling intervals at peak power achieved during the ramp-incremental exercise test in visit 1 interspersed with 75 seconds of low-intensity (30 W) cycling (Figure 1b). Heart rate, VO_2_, and the respiratory exchange ratio (RER) were recorded during the fifth exercise interval. Training consisted of nine training sessions and used the same protocol as in the acute exercise trial on non-consecutive days over 3 weeks. Training volume increased from 8 to 12 intervals over the nine training sessions while the power at which participants cycled during the high-intensity intervals remained the same throughout. Participants completed a post-training 20 km time trial 48 hours after the final training session and then returned 48 hours after this time trial to provide a muscle biopsy and blood sample and complete a post-training VO_2peak_ test.

**Figure 1.**
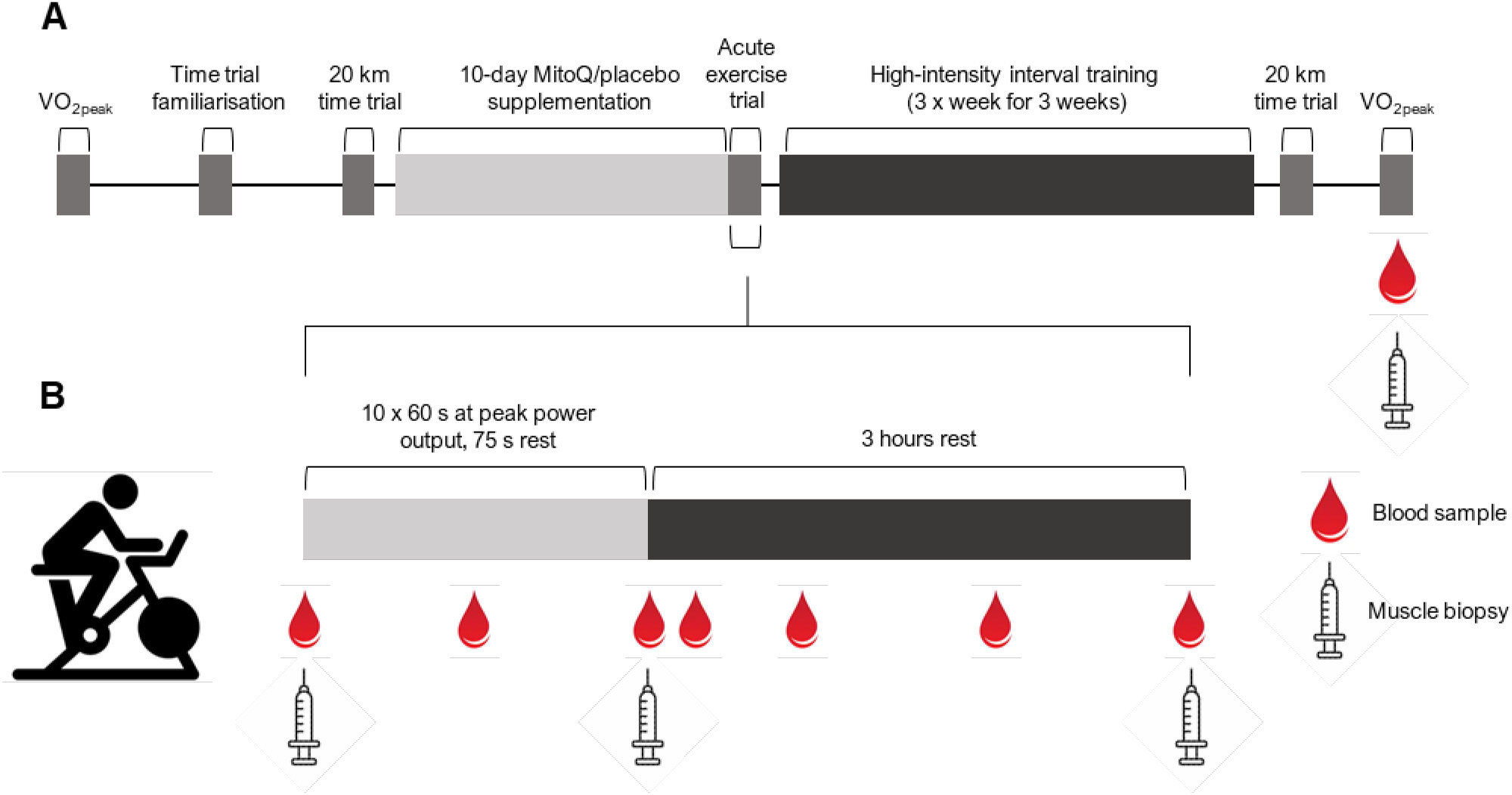
Study timeline and acute exercise trial procedures. Participants completed a VO_2peak_ test and 20 km time trial before supplementing with MitoQ or a placebo for 10 days. Participants then completed a bout of high-intensity interval exercise (10 × 60 s at VO_2peak_ power output). Muscle biopsies were collected before exercise, immediately after exercise, and 3 hours after exercise. Blood samples were collected before exercise and immediately, 30 min, and 1, 2, and 3 hours after exercise. Participants then completed 9 sessions of high-intensity interval training before repeating the 20 km time trial and VO_2peak_ test. A muscle biopsy and blood sample were collected on completion of training.

### Supplementation

In a double blind, placebo-controlled design, participants were randomized by an independent researcher to receive MitoQ (mitoquinone mesylate 20 mg/day, Alaron; Nelson, New Zealand) or an identical placebo (Alaron; Nelson, New Zealand [tapioca powder, precipitated silica and microcrystalline cellulose 101]). Tablets were dispensed into unmarked bottles by the independent researcher who performed the randomisation, and these bottles were given to the researcher conducting the trial for distribution to participants. Participants were instructed to consume one tablet per day 30 minutes before breakfast for 10 days before the acute exercise trial until completion of the final study visit. Adherence to the supplementation protocol was monitored by oversupplying tablets and counting the number of tablets returned at the final study visit. Participants were asked if they had experienced any side effects of supplementation at the acute exercise trial.

### Blood and muscle sampling

Blood samples were collected into K2 EDTA vacutainer tubes from a cannula inserted into an antecubital vein before exercise, midway through exercise, immediately after exercise, 15 minutes after exercise, and 1, 2, and 3 hours after exercise during the acute exercise trial and following completion of training. Whole blood samples were centrifuged at 2000 g for 10 min at 4°C and plasma was recovered and stored at -80°C until analysis. Percutaneous muscle biopsies were obtained from the vastus lateralis muscle before exercise, immediately after exercise, and 3 hours after exercise during the acute exercise trial and following completion of training using a suction-modified Bergstrom biopsy needle under local anaesthesia. Muscle samples were either placed in ice-cold histidine-tryptophan-ketoglutarate (HTK, in mM: 180 histidine, 30 mannitol, 18 histidine-HCl, 15 NaCl, 9 KCl, 4 MgCl, 2 tryptophan, 1 KH-2-oxoglutarate, 0.015 CaCl_2_) transplant buffer (Custodiol, Alsbech Hähnlein, Germany) for mitochondrial respiration analysis or snap-frozen in liquid nitrogen and stored at -80°C until analysis.

### Plasma analysis

Plasma lactate was measured using a Cobas 311 analyser (Roche, Mannheim, Germany). Plasma protein carbonyls were measured using a commercially available ELISA kit (BioCell Corporation, Auckland, New Zealand) according to the manufacturer’s instructions. Reduced glutathione (GSH) was measured in blood samples using the Bioxytech GSH/GSSG-412 assay kit (OxisResearch, Portland, OR, USA) according to the manufacturer’s instructions.

### Skeletal muscle mtDNA damage

A Long Amplicon-Quantitative PCR (LA-qPCR) assay was used to quantify total mtDNA damage in skeletal muscle samples as described previously [26]. DNA was extracted from muscle using the AllPrep DNA/RNA/miRNA Universal kit (Qiagen, Hilden, Germany) according to manufacturer’s instructions. DNA quality and purity were quantified using a Nanodrop One (Thermo Fisher Scientific, Rockford, IL) spectrometry method (A_260_/A_280_ > 1.8). DNA was quantified using PicoGreen (Thermo Fisher Scientific, Rockford, IL) according to the manufacturer’s instructions. Fluorescence was measured with a 485 nm excitation filter and a 530 nm emission filter and Lambda DNA/HindIII was used to construct a standard curve to determine the DNA concentration of unknown samples.

PCR reactions were prepared by combining the following: 10 μl Platinum SuperFi II PCR Master Mix, 5 μl 3 ng/μl sample DNA (total 15 ng template), 0.5 μl of each 10 μM primer and Nuclease-free H_2_O for a final volume of 25 μl. Primer sequences are outlined in Supplementary Table 1. Each experiment included a no template control and a 50% control, which contained control DNA. These control samples ensured a lack of contamination in the reaction components and ensured that fragment amplification remained within the linear range. Each sample was vortexed briefly and subsequently centrifuged for 10 seconds before being amplified in a thermal cycler based on the conditions presented in Supplementary Table 2.

To quantify products, PCR products were diluted 1 in 10 in TE buffer and 100 μl of the diluted PCR product was added to a white 96-well plate. 100 μl of the PicoGreen reagent was added to each well and the plate was incubated in the dark at room temperature for 5 minutes. Fluorescence was measured with an excitation of 485 nm and emission of 530 nm. To control for differences in mitochondrial content between samples, large mitochondrial PCR products were normalised for copy number using fluorescence values of small mitochondrial PCR products as previously described [27].

### Gene expression analyses

RNA was extracted from muscle using the AllPrep DNA/RNA/miRNA Universal kit (Qiagen, Hilden, Germany) according to the manufacturer’s instructions. The quantity and purity of extracted RNA was quantified by spectrophotometry (Nanodrop One; Thermo Fisher Scientific, Rockford, IL). The Nanostring nCounter gene expression assay (Nanostring Technologies, Seattle, WA) was used to quantify the expression of mitochondrial and antioxidant genes as previously described [28]. Briefly, customised probes carrying a unique fluorescent bar code were hybridized to 70 ng of RNA at 67°C for >16 hours. Purification and binding to the imaging membrane was performed using the nCounter prep station, after which the expression level of each target gene of interest was measured by counting the number of times the relevant barcode was detected using the nCounter Digital Analyzer. Analyses of counts was performed using nSolver 4.0 and mRNA counts were normalised using a pooled reference sample and the geometric mean of four housekeeping genes (HPRT1, EMC7, C1orf43, VCP). Probe sequences are given in Supplementary Table 1.

Quantitative real-time PCR (qRT-PCR) was used to assess the effect of MitoQ on exercise-induced skeletal muscle VEGF mRNA expression as hypotheses formed after the Nanostring nCounter gene expression assay had been completed. RNA used for qRT-PCR was reverse-transcribed using a High-Capacity RNA-to-cDNA kit (Life Technologies, Carlsbad, CA). Quantification of genes of interest was performed using a QuantStudio 6 PCR System using SYBR green select master mix (Applied Biosystems, Foster City, CA). Each sample was loaded in triplicate and normalized to the geometric mean of two reference genes (B2M and 36B4) Genes of interest were expressed using the delta-delta Ct (ΔΔCt) method. Primer sequences are given in Supplementary Table 1.

### Protein expression analyses and immunoblotting

Muscle protein samples were extracted with modified RIPA buffer (50 mM Tris, pH 8.0, 75 mM NaCl, 0.3% NP-40, 1% sodium deoxycholate, 0.1% SDS) with EDTA-free protease and phosphatase inhibitors (Sigma Aldrich, St Louis, MO). Protein concentration was determined by BCA assay and sample concentrations were adjusted with Laemmli buffer. Samples were heated at 95°C for 5 minutes. Proteins were separated in polyacrylamide gels and transferred to nitrocellulose membranes (Bio-Rad, Hercules, CA) using a semidry transfer system (Transblot Turbo, Bio-Rad, Hercules, CA). Membranes were blocked in 2% fish gelatin (Sigma Aldrich, St Louis, MO) and then incubated with primary antibody overnight at 4°C [GAPDH (Abcam ab9485; 1:10,000), phosphorylated p38 Thr180/Tyr182 (Cell Signalling Technology 4511; 1:1000), total p38 (Cell Signalling Technology 9212; 1:1000), phosphorylated AMPK Thr172 (Cell Signalling Technology 2531; 1:500), total AMPK (Cell Signalling 2532S; 1:5,000), VEGF (Abcam ab46154; 1:2000)]. The following day membranes were incubated with HRP-conjugated secondary antibodies (1:10,000) for 1 hour at room temperature and then developed and imaged using Clarity Western ECL (Bio-Rad, Hercules, CA) chemiluminescence and Chemidoc XRS+ system (Bio-Rad, Hercules, CA). Band intensities were quantified using ImageJ software and each band was normalized to a gel control sample in each respective gel and to a loading control protein.

### Citrate synthase enzymatic activity

Citrate synthase (CS) activity was determined in skeletal muscle samples collected before and after training using the same protein extract. Absorbance was measured at 412 nm on a plate reader (BioTek, Winooski, VT) at 25°C in the presence of 0.5 mM oxaloacetate in 50 mM Tris HCl buffer pH 8 containing 0.1 mM acetyl CoA and 0.2 mM 5,5′-dithiobis-2-nitrobenzoic acid (DTNB). The slope of CS absorbance change was calculated using an extinction coefficient of 13.6 mM^−1^ cm^−1^ and normalised to protein content.

### Mitochondrial respiration

Mitochondrial respiration was measured in fresh skeletal muscle biopsies collected before and after training as previously described [29]. Skeletal muscle fibres were separated in ice-cold BIOPS buffer (in mM: 50 K-MES, 30 sucrose, 20 taurine, 20 imidazole, 15 PCr, 7.23 K_2_EGTA, 2.77 CaK_2_EGTA, 6.56 MgCl_2_, 5.7 ATP, and 0.5 dithiothreitol, pH 7.1 at 0°C) and permeabilised with 50 ug/ml saponin for 30 minutes on ice. Following permeabilization, fibres were washed 3 × 10 minutes in ice-cold MiRO5 buffer (in mM: 110 sucrose, 60 K-lactobionate, 20 HEPES, 20 taurine, 10 KH_2_PO_4_, 3 MgCl_2_, 0.5 EGTA, and 1 mg/ml fraction V BSA, pH 7.1 at 37°C) and dry weight was recorded.

O_2_ consumption rate was measured in permeabilised fibres via high resolution respirometry (Oroboros Instruments, Innsbruck, Austria). Approximately 2-3 mg of tissue was added to each chamber and O_2_ concentration was maintained above 300 μM to overcome O_2_ diffusion barriers in the fibres. Malate (2 mM) and pyruvate (5 mM) were added to initiate complex I (CI) leak respiration. Succinate (10 mM) was then added to measure CI and CII leak respiration after which ADP was titrated to initiate CI and CII oxidative phosphorylation (OXPHOS). Oligomycin (2.5 μM) was added to inhibit ATP synthase and CI and CII leak respiration was measured again allowing for the calculation of leak oxygen consumption and H_2_O_2_ in the presence of ADP without ATP synthesis. Uncoupled respiration was measured following the addition of carbonyl cyanide m-chlorophenyl hydrazine (CCCP, 0.5 μM). Antimycin A (1 μM) was added to determine non-mitochondrial O_2_ consumption rate. CIV (cytochrome C oxidase, CCO) respiration was measured following the addition of ascorbate (2 mM) and N,N,N′,N′-tetramethyl-p-phenylenediamine dihydrochloride (TMPD; 0.5 mM). Finally, CIV was inhibited by the addition of azide (100 mM). All respiration rates were corrected for non-mitochondrial O_2_ consumption, CCO respiration was corrected with CIV inhibition and data are presented relative to muscle mass. All data were recorded and analysed using DatLab 7.1 software (Orobos Instruments, Innsbruck, Austria).

### Statistical analysis

All data are presented as mean ± SD unless otherwise stated. Statistical analyses were conducted in Prism 8 (GraphPad Software, Version 8) using unpaired two-tailed Student’s t-test and two-way repeated-measures ANOVA with Fisher’s least significant difference (LSD) post hoc analysis as indicated in figure legends. Statistical significance was determined as p<0.05.

## RESULTS

### Supplementation adherence and exercise intensity during the acute exercise trial

Adherence to supplementation instructions was >95% for both groups and there was no difference in supplementation adherence between groups (placebo; 98 ± 2%, MitoQ; 97 ± 5%, p=0.610). No side effects of supplementation were reported. MitoQ and placebo groups were matched for workload during the acute exercise bout (Table 2). Consistent with both groups experiencing similar levels of exercise stress, mean heart rate and VO_2_ recorded during the fifth interval of the acute exercise bout and plasma lactate measured immediately after exercise were similar between groups (Table 2). Despite participants exercising at >80% VO_2peak_ and having an ∼11-fold increase in plasma lactate from rest, exercise did not result in increased phosphorylation of skeletal muscle p38 MAPK or AMPK (Supplementary Figure 2). However, MitoQ supplementation was associated with higher p38 MAPK but not AMPK phosphorylation (Supplementary Figure 1).

**Table 2.** Measures of exercise intensity during the acute exercise trial.

|  | Placebo | MitoQ | p-value |
| --- | --- | --- | --- |
| Heart rate (bpm) | 155 ± 23 | 154 ± 18 | 0.929 |
| V̇O<sub>2</sub> (ml/kg/min) | 33.4 ± 6.2 | 29.5 ± 4.8 | 0.121 |
| Percentage of VO<sub>2peak</sub> (%) | 86 ± 13 | 81 ± 10 | 0.391 |
| Workload across work intervals | 237 ± 46 | 232 ± 34 | 0.825 |
| Post-exercise plasma lactate (mmol/l) | 15.16 ± 2.33 | 13.02 ± 3.21 | 0.089 |

Values are mean ± SD

### Effect of MitoQ supplementation and exercise on plasma and skeletal muscle oxidative stress markers

To assess whether high-intensity interval exercise or MitoQ supplementation in healthy middle-aged men alters circulating or skeletal muscle markers of oxidative stress, we assessed plasma protein carbonyl concentration and skeletal muscle mtDNA damage. Following 10 days of MitoQ or placebo supplementation, both groups had similar resting plasma protein carbonyl concentrations (Figure 2a) and mtDNA damage (Figure 2c). Acute exercise had no effect on plasma protein carbonyl concentrations (Figure 2b) and mtDNA damage (Figure 2d). Similarly, blood reduced glutathione concentrations were not affected by MitoQ supplementation (Figure 2e) or acute exercise (Figure 2f).

**Figure 2.**
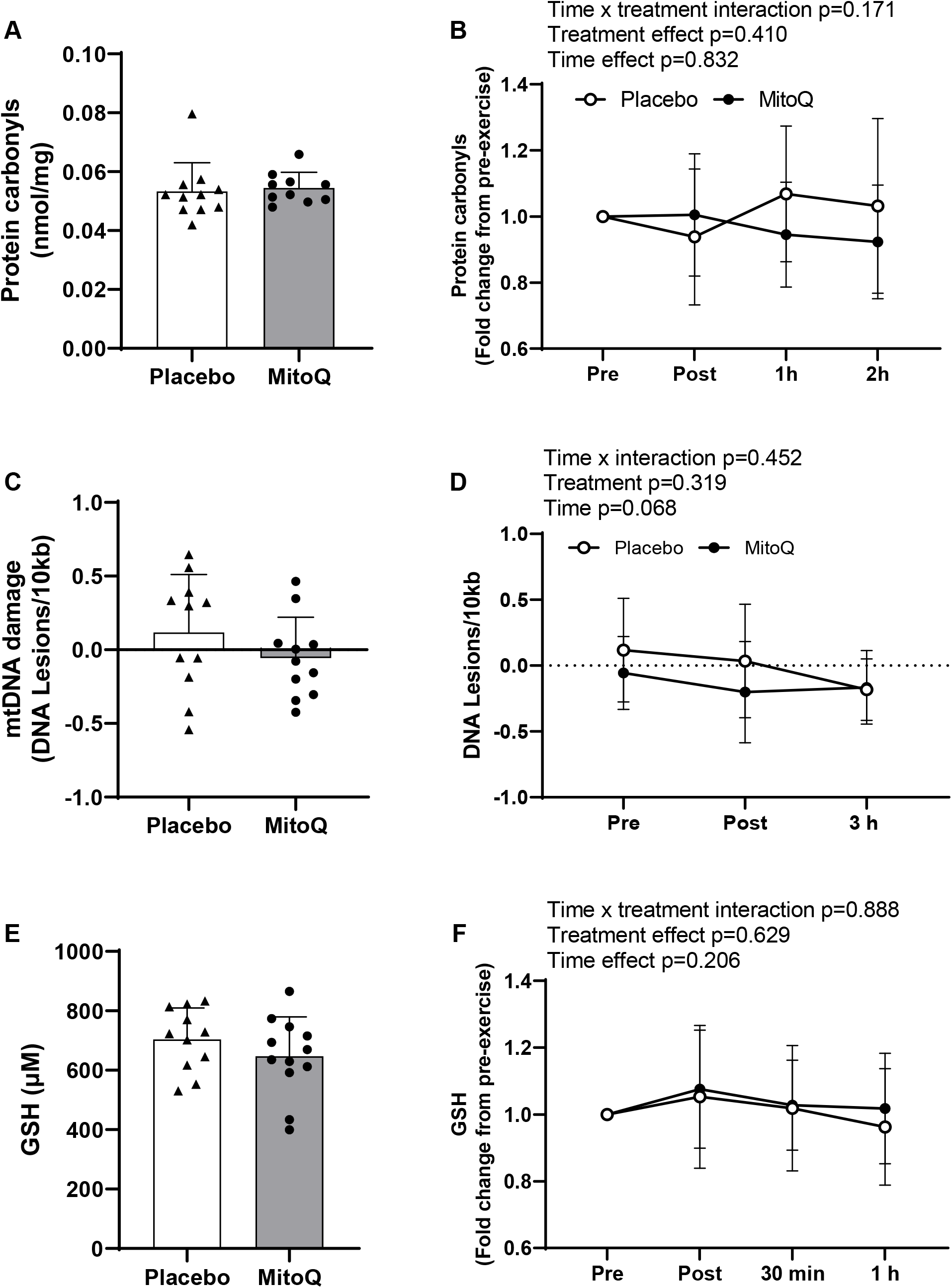
The effect of MitoQ supplementation and exercise on systemic and skeletal muscle markers of oxidative stress. A) Baseline plasma protein carbonyls following 10 days of MitoQ or placebo supplementation. B) Plasma protein carbonyls immediately (Post), 1 hour (1 h), and 2 hours (2 h) after exercise presented as fold change from pre-exercise. C) Baseline skeletal muscle mtDNA damage following 10 days of MitoQ or placebo supplementation. D) Skeletal muscle mtDNA damage immediately and 3 hours (3 h) after exercise. E) Baseline blood reduced glutathione (GSH) concentration following 10 days of MitoQ or placebo supplementation. F) Blood GSH concentration immediately, 30 min, and 1 h after exercise presented as fold change from pre-exercise. Triangles and circles represent individual values. Differences in markers of oxidative stress at baseline were analysed using unpaired Student’s t-test. Main two-way repeated measures ANOVA effects are given in figures B, D, and F. Data are presented as mean ± SD.

### Effect of MitoQ supplementation on the skeletal muscle transcriptional response to acute exercise

General antioxidant supplementation has been shown to attenuate acute exercise-induced increases in skeletal muscle mitochondrial biogenesis and antioxidant markers in young men [5]. PGC1*_α_* is a major regulator of proteins involved in exercise-induced mitochondrial biogenesis and antioxidant enzymes, therefore, we assessed *PPARGC1A* mRNA levels in skeletal muscle immediately (post) and 3 h after exercise. *PPARGC1A* mRNA was significantly increased 3 h after exercise, and this exercise-induced increase was greater in the MitoQ group compared to placebo (placebo; 3.15 ± 2.50-fold, MitoQ; 6.80 ± 3.42-fold, interaction effect p=0.008; Figure 3a). Surprisingly, in this middle-aged cohort, acute high-intensity exercise did not affect expression of the mitochondrial biogenesis marker *COX4* mRNA, and resulted in a decrease in the expression of *TFAM* and *CYTB* mRNA, which was not affected by MitoQ supplementation (Figure 3, b-d). MitoQ supplementation did not affect the basal (resting) mRNA expression of assessed mitochondrial and antioxidant markers (Figure 3e). Neither MitoQ or exercise altered the expression of the cytosolic antioxidant genes *CAT* or *TRX1* while *GPX1* and *SOD1* mRNA expression decreased with exercise independent of supplementation group (Figure 3f-i). In contrast, *SOD2* mRNA was increased by acute exercise and this was unaffected by MitoQ supplementation (Figure 3j).

**Figure 3.**
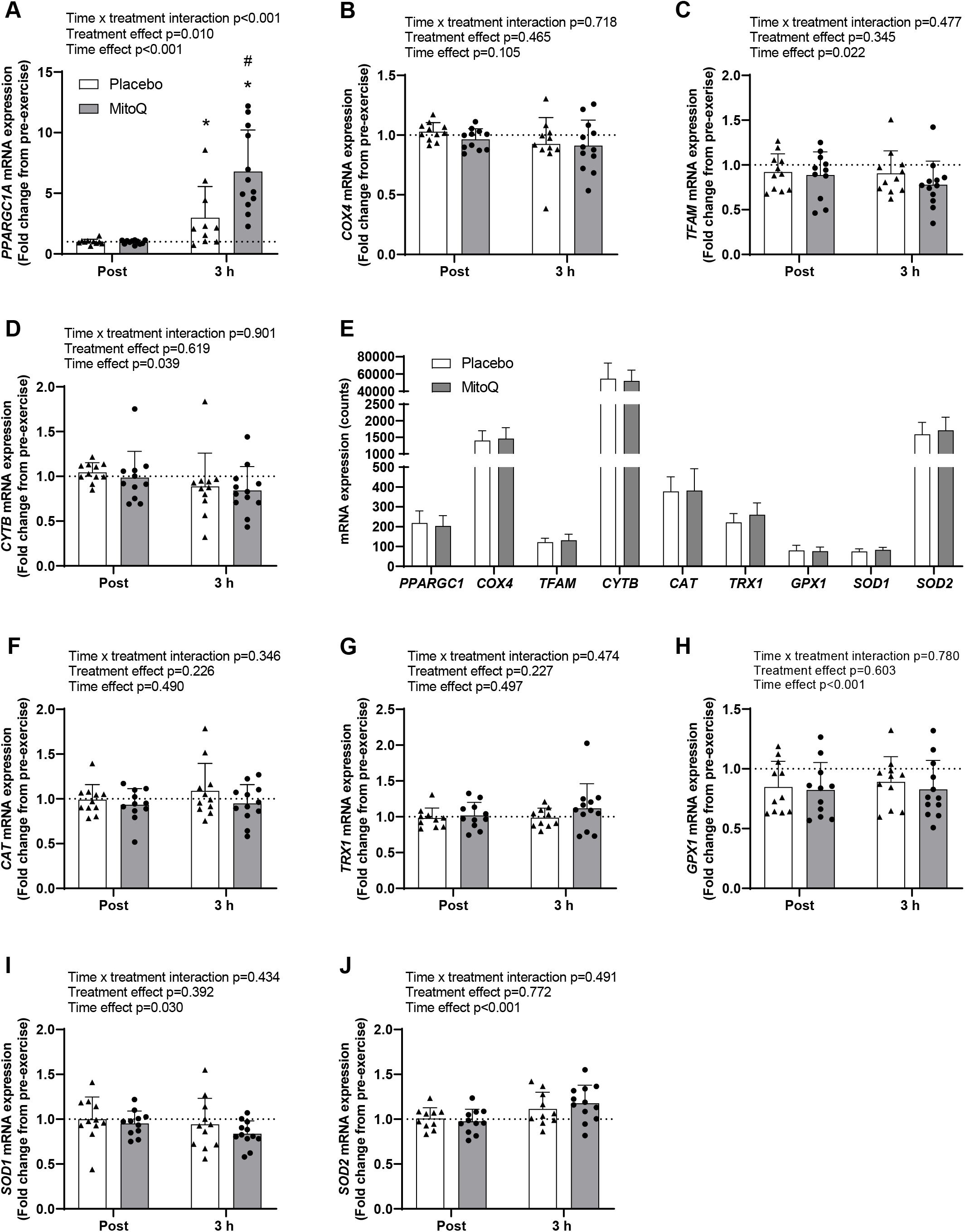
The effect of MitoQ supplementation on acute exercise-induced mitochondrial and antioxidant gene expression in skeletal muscle. A-D) Skeletal muscle mitochondrial gene expression immediately (Post) and 3 hours (3 h) after exercise following 10 days of MitoQ or placebo supplementation presented as fold change from pre-exercise. E) Baseline skeletal muscle mitochondrial and antioxidant gene expression following 10 days of MitoQ or placebo supplementation. F-J) Skeletal muscle antioxidant gene expression immediately and 3 hours after exercise following 10 days of MitoQ or placebo supplementation presented as fold change from pre-exercise Triangles and circles represent individual values. Differences in skeletal muscle mitochondrial and antioxidant gene expression at baseline were analysed using unpaired Student’s t-test. Main two-way repeated measures ANOVA effects are given in figures A-D and F-J. * p<0.05 for Fisher’s LSD post hoc vs. Pre of the same group and # p<0.05 for Fisher’s LSD post hoc vs. placebo group at same time point. Data are presented as mean ± SD. *PPARGC1A*; Peroxisome Proliferator-Activated Receptor Gamma Coactivator 1-Alpha, *COX4*; Cytochrome c oxidase subunit 4, *TFAM*; Mitochondrial Transcription Factor A, *CYTB*; Cytochrome B, *CAT*; Catalase, *TRX1*; Thioredoxin 1; *GPX1*; Glutathione peroxidase 1, *SOD1*; Superoxide Dismutase 1, *SOD2*; Superoxide Dismutase 2.

### Effect of MitoQ supplementation on training responses

Having observed that MitoQ supplementation augmented acute exercise-induced increases in skeletal muscle *PPARGC1A* mRNA, we next investigated whether this might translate to differential increases in performance or mitochondrial function in response to three weeks of high intensity interval training (HIIT). While MitoQ supplementation did not affect HIIT-induced improvements in VO_2peak_ (placebo; 9.52 ± 9.21%, MitoQ; 10.03 ± 11.97%, p=0.911, Figure 4a-b) or time to complete a 20 km cycling time trial (placebo; -2.36 ± 3.91 min, MitoQ; -4.43 ± 3.68 min, p=0.216 for interaction effect; Figure 4c-d), a significant interaction (time x treatment) (p=0.03) was observed for peak power achieved during the VO_2peak_ test (Figure 4e) with MitoQ supplementation resulting in a significantly greater relative increase (placebo; 8.15 ± 7.01 W, MitoQ; 13.64 ± 4.75 W, p=0.044, Figure 4f).

**Figure 4.**
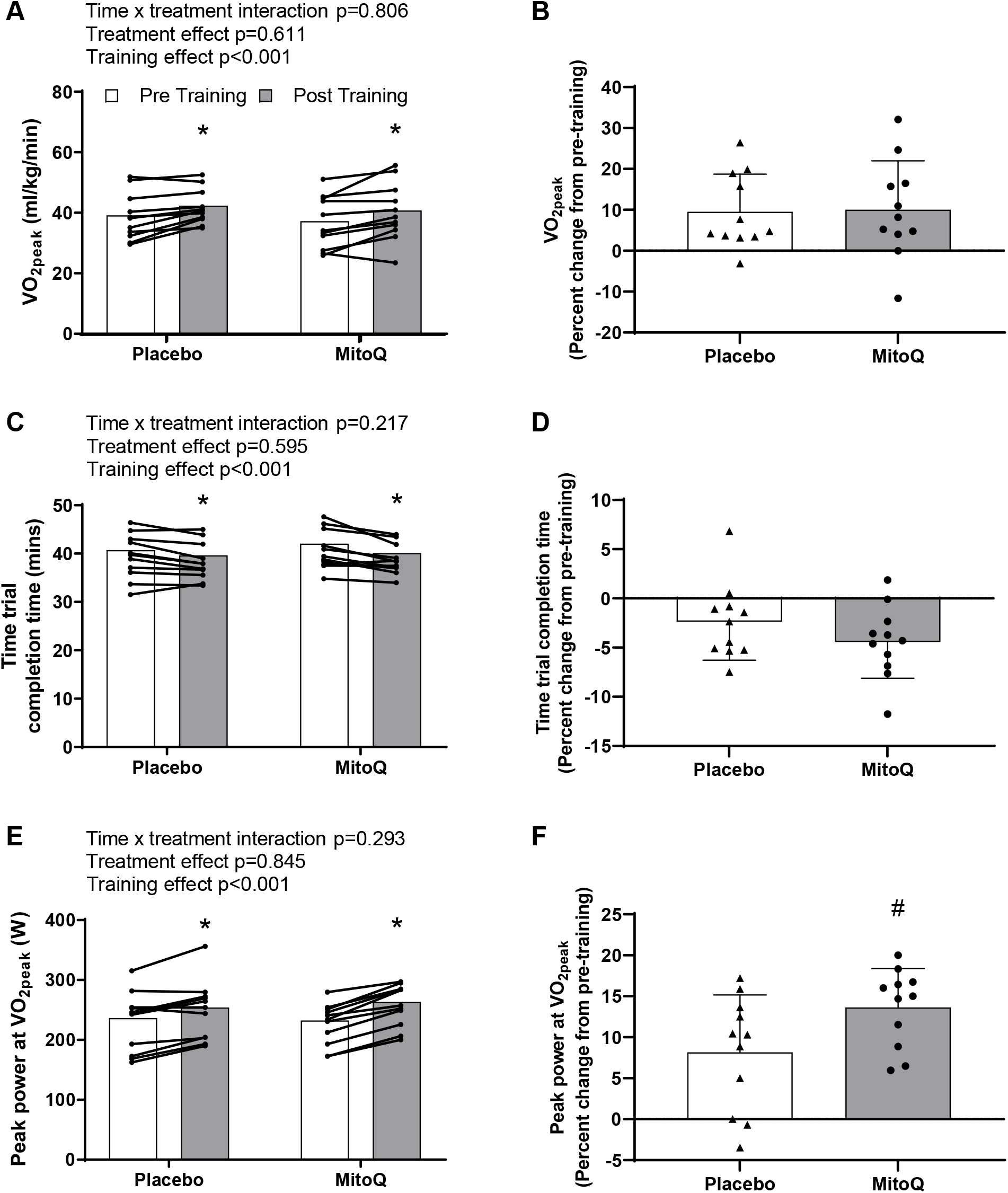
The effect of MitoQ supplementation on training-induced improvements in cycling performance. Training-induced improvements in A-B) VO_2peak_, C-D) time to complete the 20 km time trial, and E-F) peak power during the VO_2peak_ test. Triangles and circles represent individual values. Main two-way repeated measures ANOVA effects are shown in figures A, C, and E. * p<0.05 for Fisher’s LSD post hoc vs. Pre of the same group, # p<0.05 for Unpaired Student’s t-test. Data are presented as mean ± SD.

To determine whether the improvement in peak power at VO_2peak_ may be attributed to differential mitochondrial responses to exercise when supplementing with MitoQ, we assessed skeletal muscle mitochondrial content and function in skeletal muscle samples collected before and following HIIT. MitoQ and placebo supplementation groups had similar muscle citrate synthase (CS) activity prior to training (placebo; 36.06 ± 21.46 U/g protein, MitoQ; 46.97 ± 15.25 U/g protein, p=0.217) and HIIT increased CS activity to a similar extent in both groups (Figure 5a; p=0.03 for time effect). In contrast to our previous observations in young participants [30], and despite increases in CS, HIIT did not affect isolated mitochondrial oxygen consumption in any of the assessed respiration states (Figure 5b). MitoQ and placebo supplemented groups had similar mitochondrial respiration both pre- and post-HIIT. Similarly, neither HIIT or MitoQ affected basal skeletal muscle *PPARGC1A, TFAM, COX4* or *CYTB* mRNA expression (Figure 5c-d).

**Figure 5.**
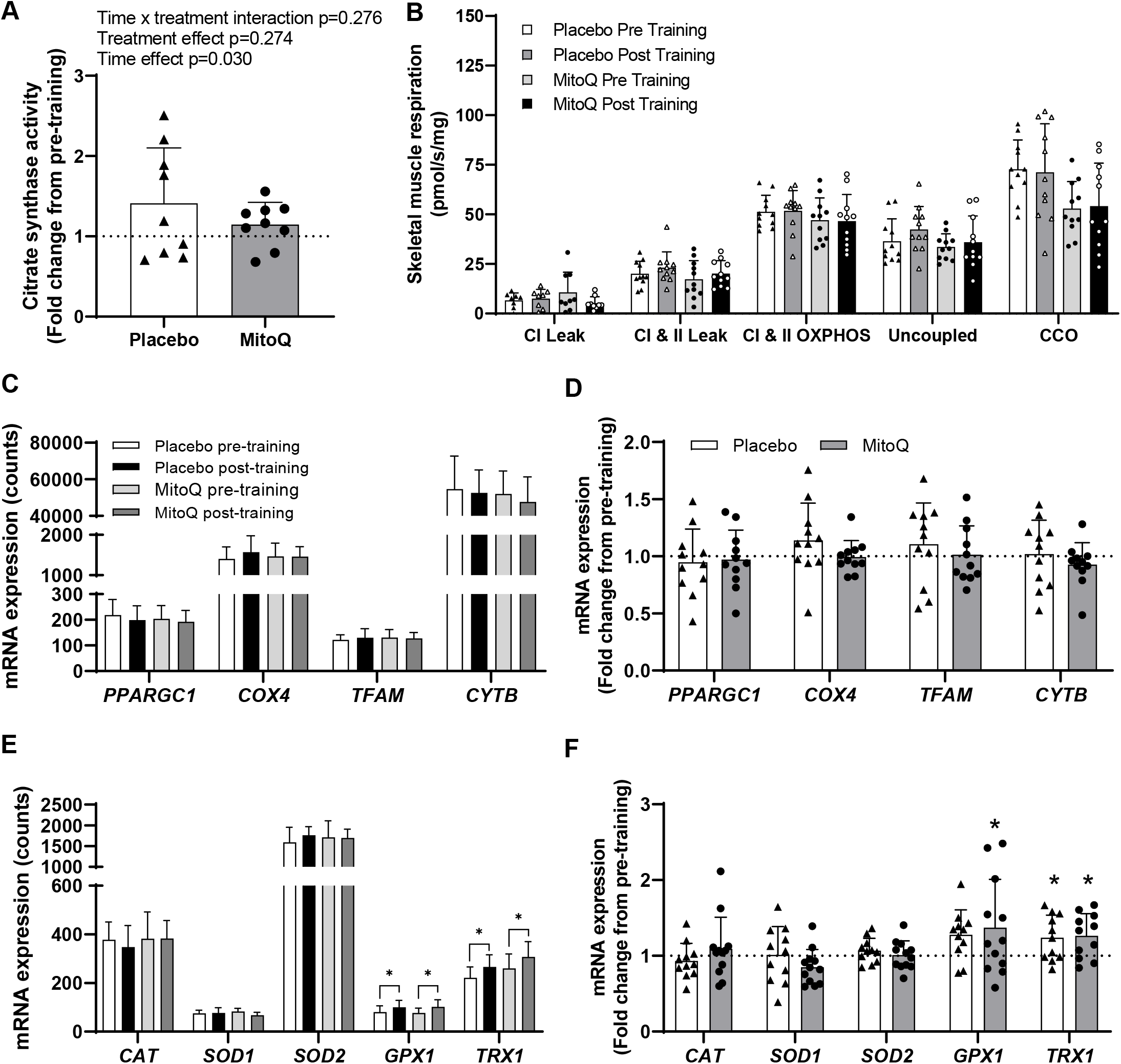
The effect of MitoQ supplementation on markers of mitochondrial content and function and antioxidant enzyme expression in skeletal muscle following HIIT. A) Skeletal muscle citrate synthase activity following 3 weeks of HIIT presented as fold change from pre-exercise. B) Skeletal muscle mitochondrial respiration before and after 3 weeks of HIIT. C) Skeletal muscle mitochondrial gene expression before and after 3 weeks of HIIT. D) Skeletal muscle mitochondrial gene expression following 3 weeks of HIIT presented as fold change from pre-exercise. E) Skeletal muscle antioxidant gene expression before and after 3 weeks of training. F) Skeletal muscle antioxidant gene expression following 3 weeks of training presented as fold change from pre-exercise Triangles and circles represent individual values. The effect of training and MitoQ supplementation on markers of skeletal muscle mitochondrial biogenesis was measured using two-way repeated measures ANOVA. Data are presented as mean ± SD. *PPARGC1A*; Peroxisome Proliferator-Activated Receptor Gamma Coactivator 1-Alpha, *COX4*; Cytochrome c oxidase subunit 4, *TFAM*; Mitochondrial Transcription Factor A, *CYTB*; Cytochrome B, *CAT*; Catalase, *TRX1*; Thioredoxin 1; *GPX1*; Glutathione peroxidase 1, *SOD1*; Superoxide Dismutase 1, *SOD2*; Superoxide Dismutase 2.

Since PGC1α also regulates expression of antioxidant enzymes [16], we measured the expression of antioxidant genes in muscle samples collected before and after training. While neither HIIT or MitoQ supplementation affected muscle *CAT, SOD1*, or *SOD2* mRNA expression (Figure 5e-f), the mRNA expression *GPX1* and *TRX1* were elevated following HIIT independent of MitoQ supplementation (Figure 5e-f).

Since MitoQ supplementation had a limited impact on post-HIIT mitochondrial function and antioxidant gene expression were next explored alternative mechanisms through which MitoQ may augment training-induced increases in peak power at VO_2peak_. MitoQ has previously been reported improve endothelial function [31] and PGC1α can drive VEGF-mediated angiogenesis [32]. While rested muscle *VEGF* mRNA (Figure 6a) and protein expression (Figure 6b) was not different between supplementation groups either pre- or post-HIIT, the acute exercise-induced increase in *VEGF* mRNA was greater in the MitoQ supplemented group (Figure 6c). Since this study was not designed to assess the effect of MitoQ on HIIT-induced skeletal muscle capillarisation muscle was not collected for histological analysis. It was also interesting to observe that despite MitoQ augmenting peak power output during the VO_2peak_ test post-HIIT, training induced increases in VO_2peak_ were not affected. This may suggest that MitoQ enhances HIIT-associated increases in maximal anaerobic capacity, however mRNA expression of anaerobic metabolism genes *LDHA* and *HKII* was similar in skeletal muscle from MitoQ and placebo treated participants (Supplementary Figure 3).

**Figure 6.**
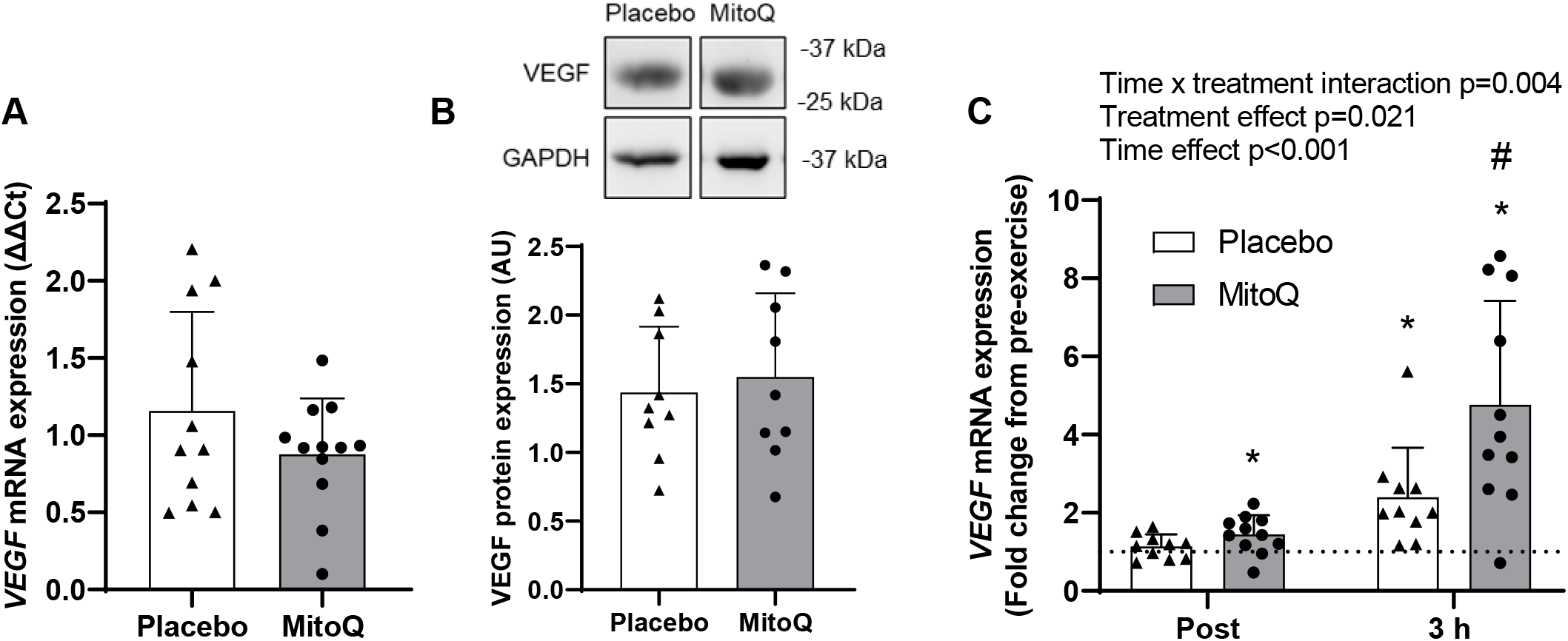
The effect of MitoQ supplementation on exercise-induced increases in skeletal muscle VEGF mRNA expression. A) Baseline skeletal muscle *VEGF* mRNA expression following 10 days of MitoQ or placebo supplementation. B) Representative blot of baseline skeletal muscle VEGF protein expression following 10 days of MitoQ or placebo supplementation and quantification of VEGF protein expression relative to GAPDH protein expression. Representative bands were run on the same gel and images have been cropped to show VEGF protein expression in participants from the placebo and MitoQ group together. C) Skeletal muscle *VEGF* gene expression immediately (Post) and 3 hours (3 h) after exercise following 10 days of MitoQ or placebo supplementation presented as fold change from pre-exercise levels. Triangles and circles represent individual values. Differences in skeletal muscle VEGF mRNA and protein expression at baseline were analysed using unpaired Student’s t-test. Main two-way repeated measures ANOVA effects are shown in figure C. * p<0.05 for Fisher’s LSD post hoc vs. Pre of the same group and # p<0.05 for Fisher’s LSD post hoc vs. placebo group at same time point. Data are presented as mean ± SD. VEGF; Vascular Endothelial Growth Factor, GAPDH; Glyceraldehyde 3-phosphate dehydrogenase.

## DISCUSSION

Supplementation with non-targeted antioxidants can result in indiscriminate neutralisation of ROS from multiple intracellular sources and has been shown to attenuate acute exercise-induced activation of redox-sensitive signalling pathways in some instances [4, 13]. Consequently, supplementing with non-targeted antioxidants during exercise training may negate or attenuate some of the major beneficial effects of exercise [33]. Here, we found that supplementation with the mitochondria-targeted antioxidant MitoQ did not attenuate HIIT-induced increases in performance (VO_2peak_ and cycling time trial) or skeletal muscle antioxidant gene expression and citrate synthase activity, which is a marker of mitochondrial content. Moreover, MitoQ supplementation was associated with greater acute high-intensity exercise-induced increases in skeletal muscle PGC1α mRNA expression and post-training improvements in peak power during VO_2peak_ assessment.

The enhancement of acute exercise-induced increases in skeletal muscle PGC1α mRNA expression in the MitoQ supplemented group may, in part, explain the greater HIIT associated improvement in peak power during the VO_2peak_ test. PGC1α is the major regulator of mitochondrial biogenesis [34]; however, despite this, our results indicate that HIIT-associated increases in skeletal muscle citrate synthase were not affected by MitoQ supplementation. This is in agreement with our findings, and others [35], that MitoQ supplementation does not affect post-exercise training VO_2peak_, skeletal muscle mitochondrial function, or oxidative capacity (as assessed by near-infrared spectroscopy), and suggests that MitoQ may be acting independent of mitochondrial adaptations to improve post-training peak power output. While it is possible this effect is PGC1α-independent, acute exercise induced increases in PGC1α have been implicated in coordinating a host of adaptive responses to exercise, including antioxidant expression [36] and angiogenesis (via VEGF) [32]. Unlike some general antioxidant supplements [11], MitoQ supplementation did not alter exercise-induced changes in antioxidant gene expression. However, acute high-intensity exercise-induced increases in muscle VEGF mRNA expression were augmented, potentially suggesting that MitoQ may enhance training-induced increases in skeletal muscle vascularisation and muscle perfusion to increase peak power output. It is possible that an increased skeletal muscle capillary density in the MitoQ group enabled greater metabolite clearance during the VO_2peak_ test, resulting in improved time to exhaustion.

We have recently shown that MitoQ supplementation improves 8 km time trial performance in trained cyclists, and this was associated with attenuated exercise-induced increases in plasma F_2_-isoprostanes [37]. Furthermore, infusion of the antioxidant N-acetylcysteine has been shown to improve performance in highly trained cyclists, who can achieve very high workloads during a performance test [10]. In the present study, MitoQ improved peak power output during the VO_2peak_ test, but had no effect on 20 km time trial performance when the participant’s power output was much lower. Taken together, these results may indicate that oxidative stress plays a more significant role in impairing performance at exercise intensities that require significant anaerobic component, and would be consistent with trained cyclists being able achieve higher power output and plasma lactate during short-duration (<15 min) all out exercise when supplemented with MitoQ [37]. It is therefore possible that MitoQ is acting as a direct ergogenic aid rather than enhancing exercise-training induced improvements in peak power output.

Unfortunately, blood and tissue samples were not collected during the VO_2peak_ tests meaning we could not assess the impact of MitoQ on oxidative stress markers during these tests. Surprisingly, there was no effect of high-intensity interval exercise on the plasma or muscle markers of oxidative stress assessed in the present study. The duration and/or intensity of the exercise protocol used in this study may have been insufficient to induce increases in plasma protein carbonyls and skeletal muscle mtDNA damage in this participant group. Increases in skeletal muscle mtDNA damage have recently been observed following exercise consisting of longer high-intensity intervals and a more intense active recovery in young (25 years) men [18]. The age of the participants may explain the failure of exercise to increase systemic and skeletal muscle markers of oxidative stress in this study. Age-associated increases in basal skeletal muscle ROS levels are associated with increased macromolecule damage [38] and, as a consequence of basal stress, the ability of aged skeletal muscle to further increase ROS production during exercise is attenuated [39]. Therefore, it is possible that participants in this study had elevated basal levels of plasma protein carbonyls and mtDNA damage, which could not be further elevated by exercise.

It remains to be determined whether augmentation of acute exercise-induced increases in PGC1α and training induced increases in peak power output by MitoQ are the result of the antioxidant properties of MitoQ or other mechanisms. The triphenylphosphonium cation of MitoQ has been shown to exert biological effects independent of the ubiquinone moiety [40]. However, it has been suggested that age-associated oxidative stress can supress the ability of cytosolic proteins to respond to acute oxidative redox signals during exercise [41]. Decreasing mitochondrial ROS (the primary source of ROS in the basal state) through MitoQ supplementation [29] may restore responsiveness of cytosolic signalling proteins to exercise-induced ROS and therefore enhance redox activation of PGC1α. Alternatively, MitoQ may augment acute exercise-induced increases in PGC1α mRNA expression through a mechanism involving hypomethylation of the PGC1α promotor region [42] as ROS generated by mitochondria can affect DNA methylation by acting on the activity and expression of DNA methyltransferases [43].

There are several limitations that should be considered when interpreting the results of this study. Participants in this study were all middle-aged untrained males, meaning caution should be taken when applying the results of this study to young trained individuals and females. The age of the participants may have resulted in a blunted response to acute exercise and exercise training in skeletal muscle. We previously reported increases in skeletal muscle mitochondrial respiration following 6 sessions of HIIT in young men [31].

While studies report similar mitochondrial adaptations in old and young individuals following longer training periods (>12 weeks) [44], it is possible that training-induced mitochondrial adaptations are blunted in aged muscle in the short term. Another limitation of this study is that we did not quantify the level of MitoQ in skeletal muscle biopsies and cannot confirm that MitoQ was present within the skeletal muscle of the participants during the acute exercise trial and exercise training.

Taken together, these results provide evidence that the acute exercise-induced transcriptional response and training-induced mitochondrial adaptations in skeletal muscle are not attenuated by MitoQ supplementation. Furthermore, MitoQ enhances the effect of exercise training on peak power achieved during a ramp-incremental exercise test, which may be related to the augmentation of skeletal muscle PGC1α mRNA expression following acute exercise by MitoQ. However, this effect does not appear to be related to an effect of MitoQ supplementation on training-induced mitochondrial biogenesis in skeletal muscle and may therefore be the result of other adaptations mediated by PGC1α.

## Data Availability

All data produced in the present study are available upon reasonable request to the authors

## Acknowledgements

We are thankful to all participants for their contribution to the study. We would also like to acknowledge Randall D’Souza and Chris Hedges for their technical support during this study. TLM is supported by a Rutherford Discovery Fellowship. SCB is supported by a Callaghan Innovation Research & Development grant.

## TABLES

**Supplementary Table 1.**
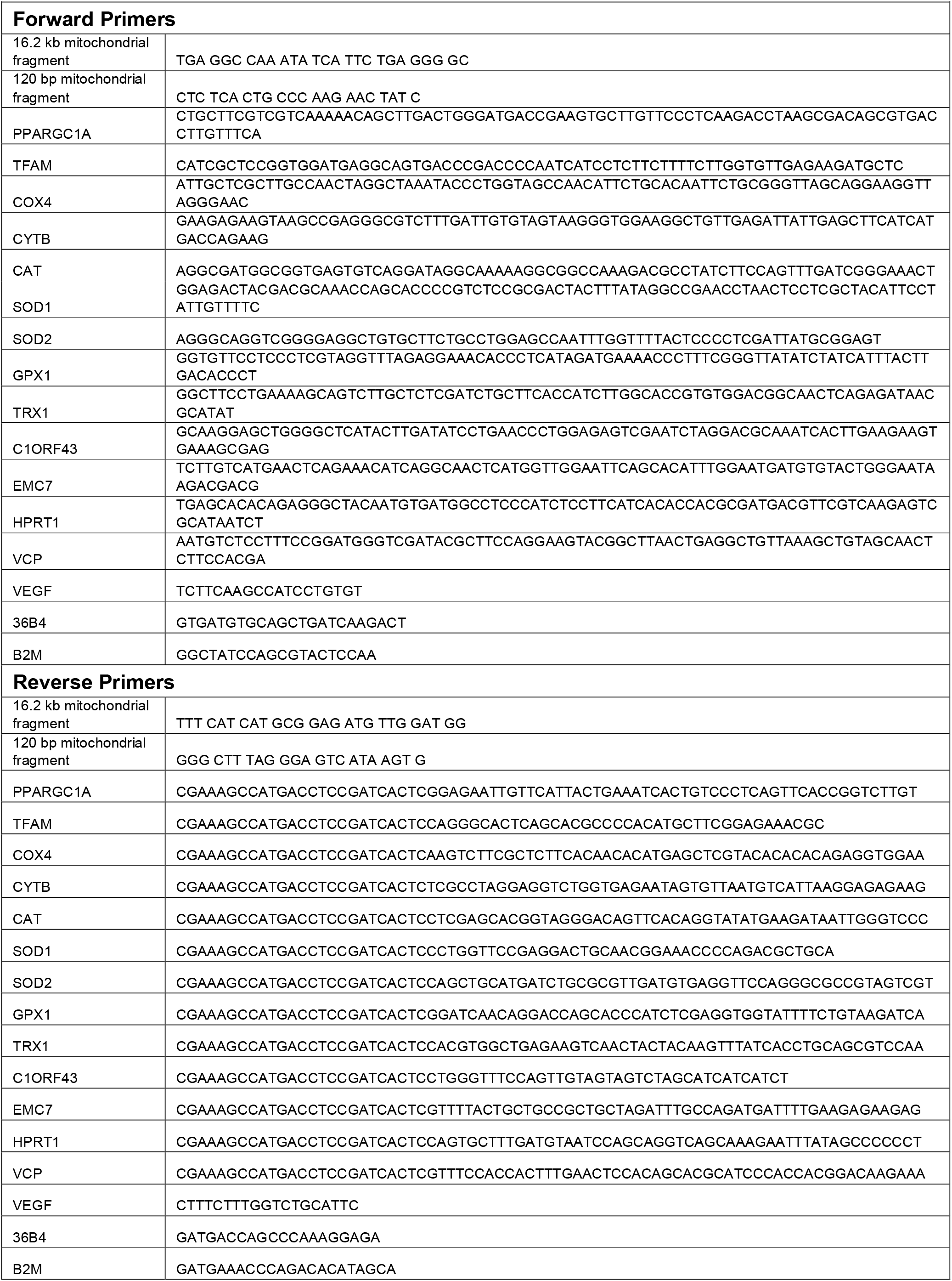
Primer sequences.

**Supplementary Table 2.**
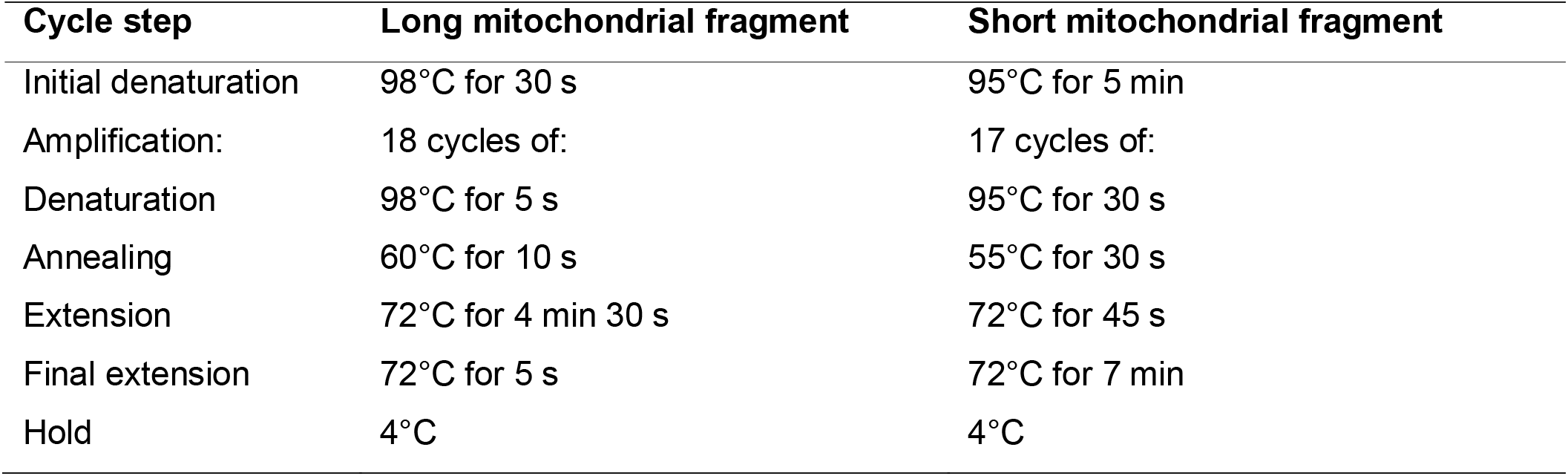
Thermal cycler conditions for amplification of long and short mitochondrial fragments.

| **Cycle step** | **Long mitochondrial fragment** | **Short mitochondrial fragment** |
| --- | --- | --- |
| Initial denaturation | 98°C for 30 s | 95°C for 5 min |
| Amplification: | 18 cycles of: | 17 cycles of: |
| Denaturation | 98°C for 5 s | 95°C for 30 s |
| Annealing | 60°C for 10 s | 55°C for 30 s |
| Extension | 72°C for 4 min 30 s | 72°C for 45 s |
| Final extension | 72°C for 5 s | 72°C for 7 min |
| Hold | 4°C | 4°C |

## FIGURE LEGENDS

**Supplementary Figure 1. CONSORT flow chart**. This figure shows the flow of patients through the trial according to the criteria recommended in the CONSORT Guidelines.

**Supplementary Figure 2. The effect of exercise and MitoQ supplementation on skeletal muscle p38 MAPK and AMPK phosphorylation**. A) Phosphorylated p38 MAPK (pp38) protein expression relative to total p38 MAPK (total p38) protein expression in skeletal muscle before exercise (Pre) and immediately (Post) and 3 hours after exercise (3 h) following 10 days of MitoQ supplementation. B) pp38 protein expression relative to total p38 protein expression immediately and 3 h after exercise presented as fold change from pre-exercise. C) Phosphorylated AMPK^Thr172^ (pAMPK ^Thr172^) protein expression relative to total AMPK protein expression in skeletal muscle before exercise and immediately and 3 hours after exercise following 10 days of MitoQ supplementation. D) pAMPK ^Thr172^ protein expression relative to total AMPK protein expression immediately and 3 h after exercise presented as fold change from pre-exercise. Triangles and circles represent individual values. Main two-way repeated-measures ANOVA effects are given in figures. Values are presented as means ± SD.

**Supplementary Figure 3. The effect of MitoQ supplementation on mRNA expression of genes involved in anaerobic metabolism**. Baseline skeletal muscle A) *LDHA* and B) *HKII* mRNA expression following 10 days of MitoQ or placebo supplementation. Skeletal muscle C) *LDHA* and D) *HKII* mRNA expression immediately (Post) and 3 hours (3 h) after exercise presented as fold change from pre-exercise. Skeletal muscle E) *LDHA* and F) *HKII* mRNA expression after 3 weeks of training presented as fold change from pre-training. Triangles and circles represent individual values. Differences in skeletal muscle *LDHA* and *HKII* mRNA expression at baseline were analysed using unpaired Student’s t-test. Main two-way repeated measures ANOVA effects are shown in figures C-F. Data are presented as mean ± SD. *LDHA*; Lactate Dehydrogenase A, *HKII*; Hexokinase II.

## REFERENCES

1. Hawley, John A., et al., Integrative Biology of Exercise. Cell, 2014. 159(4): p. 738–749.

2. Perry, C.G.R., et al., Repeated transient mRNA bursts precede increases in transcriptional and mitochondrial proteins during training in human skeletal muscle. The Journal of Physiology, 2010. 588(23): p. 4795–4810.

3. Margaritelis, N.V., et al., Redox basis of exercise physiology. Redox Biology, 2020: p. 101499.

4. Gomez-Cabrera, M.C., et al., Decreasing xanthine oxidase-mediated oxidative stress prevents useful cellular adaptations to exercise in rats. J Physiol, 2005. 567(Pt 1): p. 113–20.

5. Ristow, M., et al., Antioxidants prevent health-promoting effects of physical exercise in humans. Proc Natl Acad Sci U S A, 2009. 106(21): p. 8665–70.

6. Andrade, F.H., et al., Effect of hydrogen peroxide and dithiothreitol on contractile function of single skeletal muscle fibres from the mouse. J Physiol, 1998. 509((Pt 2)): p. 565–75.

7. D., L.G. and W. Håkan, Acute effects of reactive oxygen and nitrogen species on the contractile function of skeletal muscle. The Journal of Physiology, 2011. 589(9): p. 2119–2127.

8. Maughan, R.J., F. Depiesse, and H. Geyer, The use of dietary supplements by athletes. Journal of Sports Sciences, 2007. 25(sup1): p. S103–S113.

9. McKenna, M.J., et al., N-acetylcysteine attenuates the decline in muscle Na+,K+-pump activity and delays fatigue during prolonged exercise in humans. J Physiol, 2006. 576(Pt 1): p. 279–88.

10. Medved, I., et al., N-acetylcysteine enhances muscle cysteine and glutathione availability and attenuates fatigue during prolonged exercise in endurance-trained individuals. Journal of Applied Physiology, 2004. 97(4): p. 1477–1485.

11. Gomez-Cabrera, M.C., et al., Oral administration of vitamin C decreases muscle mitochondrial biogenesis and hampers training-induced adaptations in endurance performance. Am J Clin Nutr, 2008. 87(1): p. 142–9.

12. Strobel, N.A., et al., Antioxidant supplementation reduces skeletal muscle mitochondrial biogenesis. Med Sci Sports Exerc, 2011. 43(6): p. 1017–24.

13. Wadley, G.D., et al., Xanthine oxidase inhibition attenuates skeletal muscle signaling following acute exercise but does not impair mitochondrial adaptations to endurance training. Am J Physiol Endocrinol Metab, 2013. 304(8): p. E853–62.

14. Hood, D.A., Invited Review: Contractile activity-induced mitochondrial biogenesis in skeletal muscle. Journal of Applied Physiology, 2001. 90(3): p. 1137–1157.

15. Powers, S.K., et al., Exercise training-induced alterations in skeletal muscle antioxidant capacity: a brief review. 1999. 31(7): p. 987–997.

16. St-Pierre, J., et al., Suppression of Reactive Oxygen Species and Neurodegeneration by the PGC-1 Transcriptional Coactivators. Cell, 2006. 127(2): p. 397–408.

17. Henríquez-Olguin, C., et al., Cytosolic ROS production by NADPH oxidase 2 regulates muscle glucose uptake during exercise. Nature Communications, 2019. 10(1): p. 4623.

18. Williamson, J., et al., The Mitochondria-Targeted Antioxidant MitoQ, Attenuates Exercise-Induced Mitochondrial DNA Damage. Redox Biology, 2020: p. 101673.

19. Stretton, C., et al., 2-Cys Peroxiredoxin oxidation in response to Hydrogen Peroxide and contractile activity in skeletal muscle: A novel insight into exercise-induced redox signalling? Free Radical Biology and Medicine, 2020.

20. Petersen, A., et al., Infusion with the antioxidant N-acetylcysteine attenuates early adaptive responses to exercise in human skeletal muscle. 2012. 204(3): p. 382–392.

21. James, A.M., R.A.J. Smith, and M.P. Murphy, Antioxidant and prooxidant properties of mitochondrial Coenzyme Q. Archives of Biochemistry and Biophysics, 2004. 423(1): p. 47–56.

22. Ingold, K.U., et al., Autoxidation of lipids and antioxidation by alphatocopherol and ubiquinol in homogeneous solution and in aqueous dispersions of lipids: unrecognized consequences of lipid particle size as exemplified by oxidation of human low density lipoprotein. Proceedings of the National Academy of Sciences, 1993. 90(1): p. 45.

23. Ross, M.F., et al., Lipophilic triphenylphosphonium cations as tools in mitochondrial bioenergetics and free radical biology. Biochemistry (Mosc), 2005. 70(2): p. 222–30.

24. Murphy, M.P. and R.A. Smith, Targeting antioxidants to mitochondria by conjugation to lipophilic cations. Annu Rev Pharmacol Toxicol, 2007. 47: p. 629–56.

25. Dietary supplement use among adults : United States, 2017–2018, S. National Center for Health, Editor. 2021, http://dx.doi.org/10.15620/cdc:101131: Hyattsville, MD.

26. Santos, J.H., et al., Quantitative PCR-Based Measurement of Nuclear and Mitochondrial DNA Damage and Repair in Mammalian Cells, in DNA Repair Protocols: Mammalian Systems, D.S. Henderson, Editor. 2006, Humana Press: Totowa, NJ. p. 183–199.

27. Furda, A.M., et al., Analysis of DNA Damage and Repair in Nuclear and Mitochondrial DNA of Animal Cells Using Quantitative PCR, in DNA Repair Protocols, L. Bjergbæk, Editor. 2012, Humana Press: Totowa, NJ. p. 111–132.

28. Kulkarni, M.M., Digital Multiplexed Gene Expression Analysis Using the NanoString nCounter System. Current Protocols in Molecular Biology, 2011. 94(1): p. 25B.10.1-25B.10.17.

29. Pham, T., et al., MitoQ and CoQ10 supplementation mildly suppresses skeletal muscle mitochondrial hydrogen peroxide levels without impacting mitochondrial function in middle-aged men. 2020.

30. Hedges, C.P., et al., Peripheral blood mononuclear cells do not reflect skeletal muscle mitochondrial function or adaptation to high-intensity interval training in healthy young men. Journal of Applied Physiology, 2019. 126(2): p. 454–461.

31. Rossman, M.J., et al., Chronic Supplementation With a Mitochondrial Antioxidant (MitoQ) Improves Vascular Function in Healthy Older Adults. Hypertension, 2018.

32. Arany, Z., et al., HIF-independent regulation of VEGF and angiogenesis by the transcriptional coactivator PGC-1α. Nature, 2008. 451(7181): p. 1008–1012.

33. Merry, T.L. and M. Ristow, Do antioxidant supplements interfere with skeletal muscle adaptation to exercise training? J Physiol, 2016. 594(18): p. 5135–47.

34. Lin, J., et al., Transcriptional co-activator PGC-1α drives the formation of slow-twitch muscle fibres. Nature, 2002. 418(6899): p. 797–801.

35. Shill, D.D., et al., Mitochondria-specific antioxidant supplementation does not influence endurance exercise training-induced adaptations in circulating angiogenic cells, skeletal muscle oxidative capacity or maximal oxygen uptake. J Physiol, 2016. 594(23): p. 7005–7014.

36. Leick, L., et al., PGC-1α is required for training-induced prevention of age-associated decline in mitochondrial enzymes in mouse skeletal muscle. Experimental Gerontology, 2010. 45(5): p. 336–342.

37. Broome, S.C., et al., Mitochondria-targeted antioxidant supplementation improves 8□km time trial performance in middle-aged trained male cyclists. Journal of the International Society of Sports Nutrition, 2021. 18(1): p. 58.

38. Bailey, D.M., et al., Sedentary aging increases resting and exercise-induced intramuscular free radical formation. 2010. 109(2): p. 449–456.

39. Close, G.L., et al., Release of superoxide from skeletal muscle of adult and old mice: an experimental test of the reductive hotspot hypothesis. 2007. 6(2): p. 189–195.

40. Bond, S.T., et al., The antioxidant moiety of MitoQ imparts minimal metabolic effects in adipose tissue of high fat fed mice. 2019. 10: p. 543.

41. Jackson, M.J., On the mechanisms underlying attenuated redox responses to exercise in older individuals: A hypothesis. Free Radical Biology and Medicine, 2020. 161: p. 326–338.

42. Barrès, R., et al., Acute Exercise Remodels Promoter Methylation in Human Skeletal Muscle. Cell Metabolism, 2012. 15(3): p. 405–411.

43. Mohammed, S.A., et al., Epigenetic Control of Mitochondrial Function in the Vasculature. 2020. 7(28).

44. Cobley, J.N., et al., Exercise improves mitochondrial and redox-regulated stress responses in the elderly: better late than never! Biogerontology, 2015. 16(2): p. 249–264.

