## Supplementary figures and images for "Mitochondria-targeted antioxidant supplementation augments acute exercise-induced increases in muscle PGC1α mRNA and improves training-induced increases in peak power independent of mitochondrial content and function in untrained middle-aged men"

### Supplementary figures

**Supplementary Figure 1**

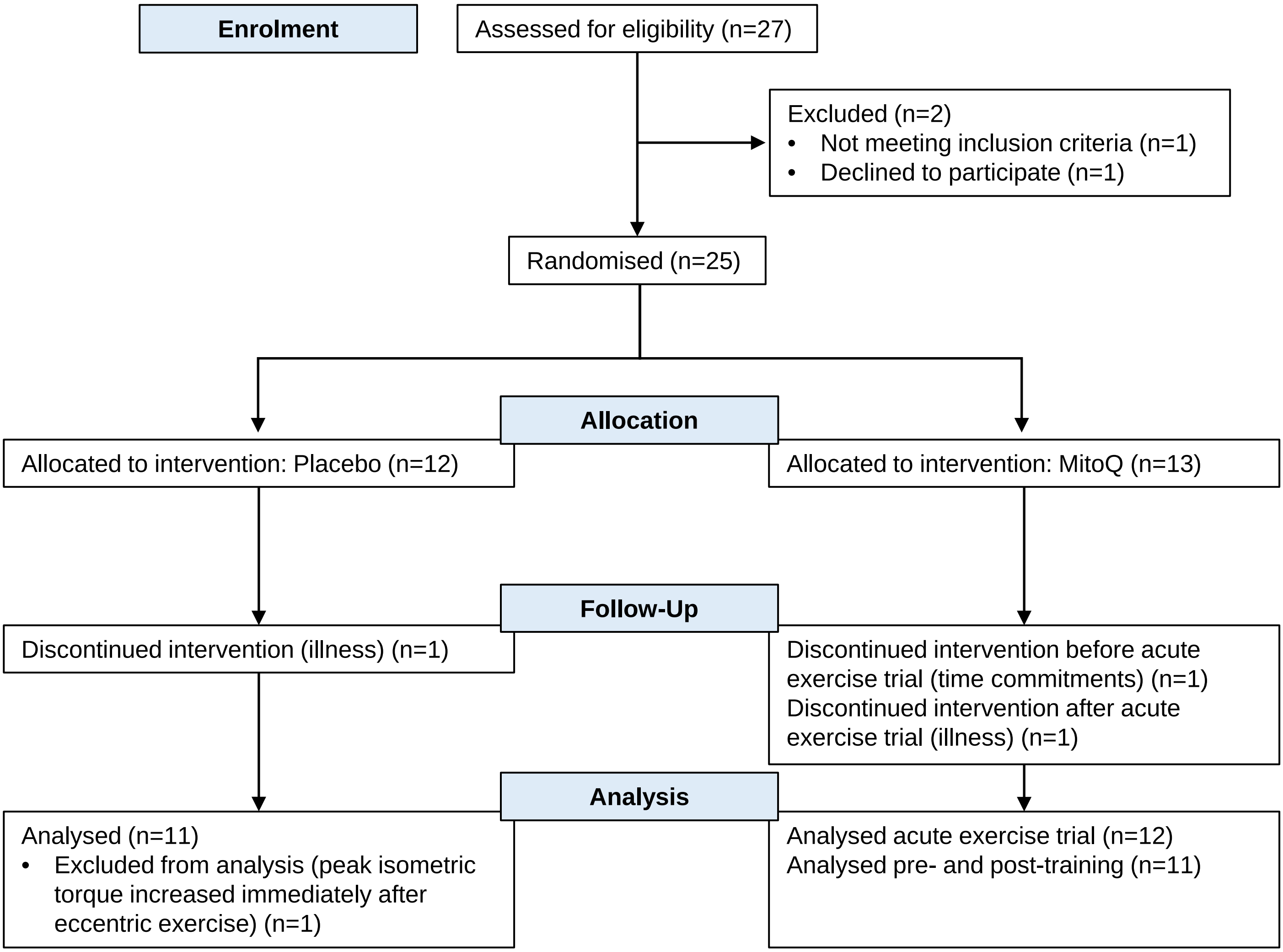

#### Supplementary Figure 2

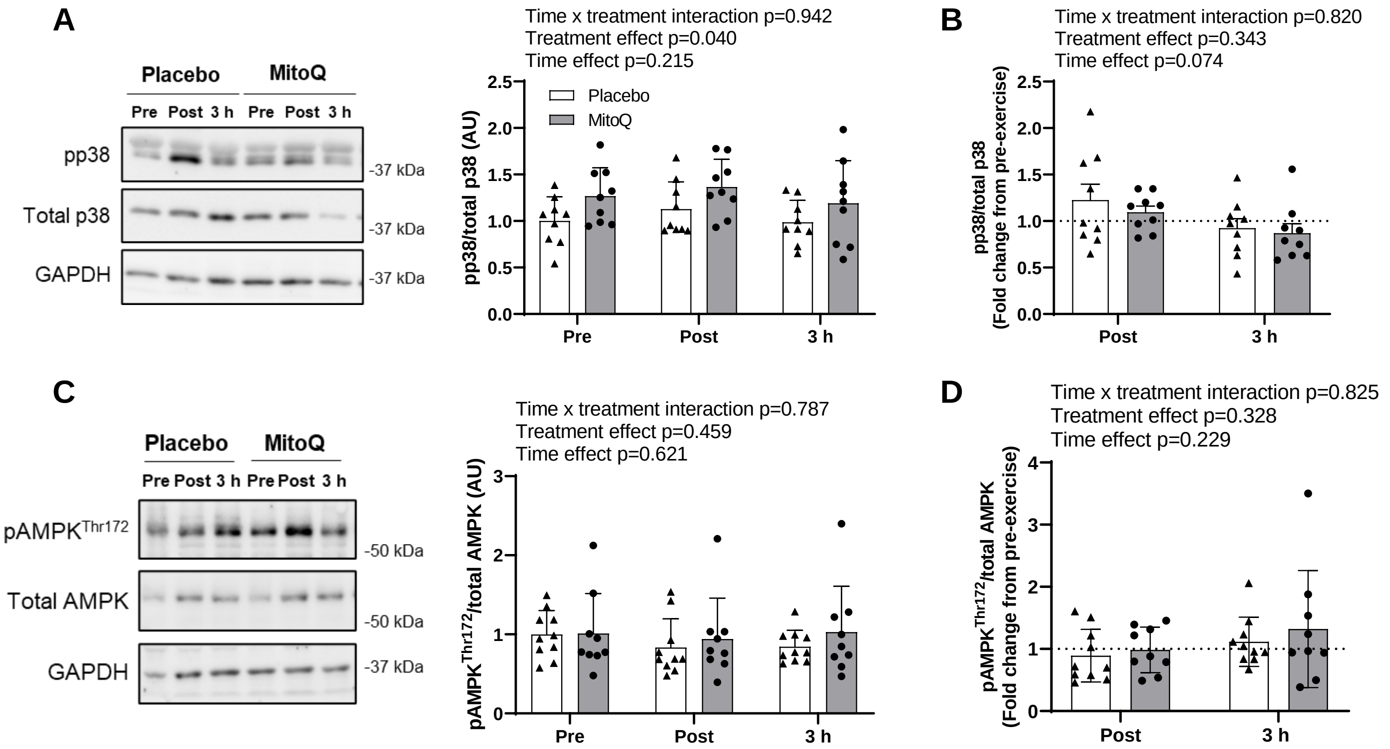

#### Supplementary Figure 3

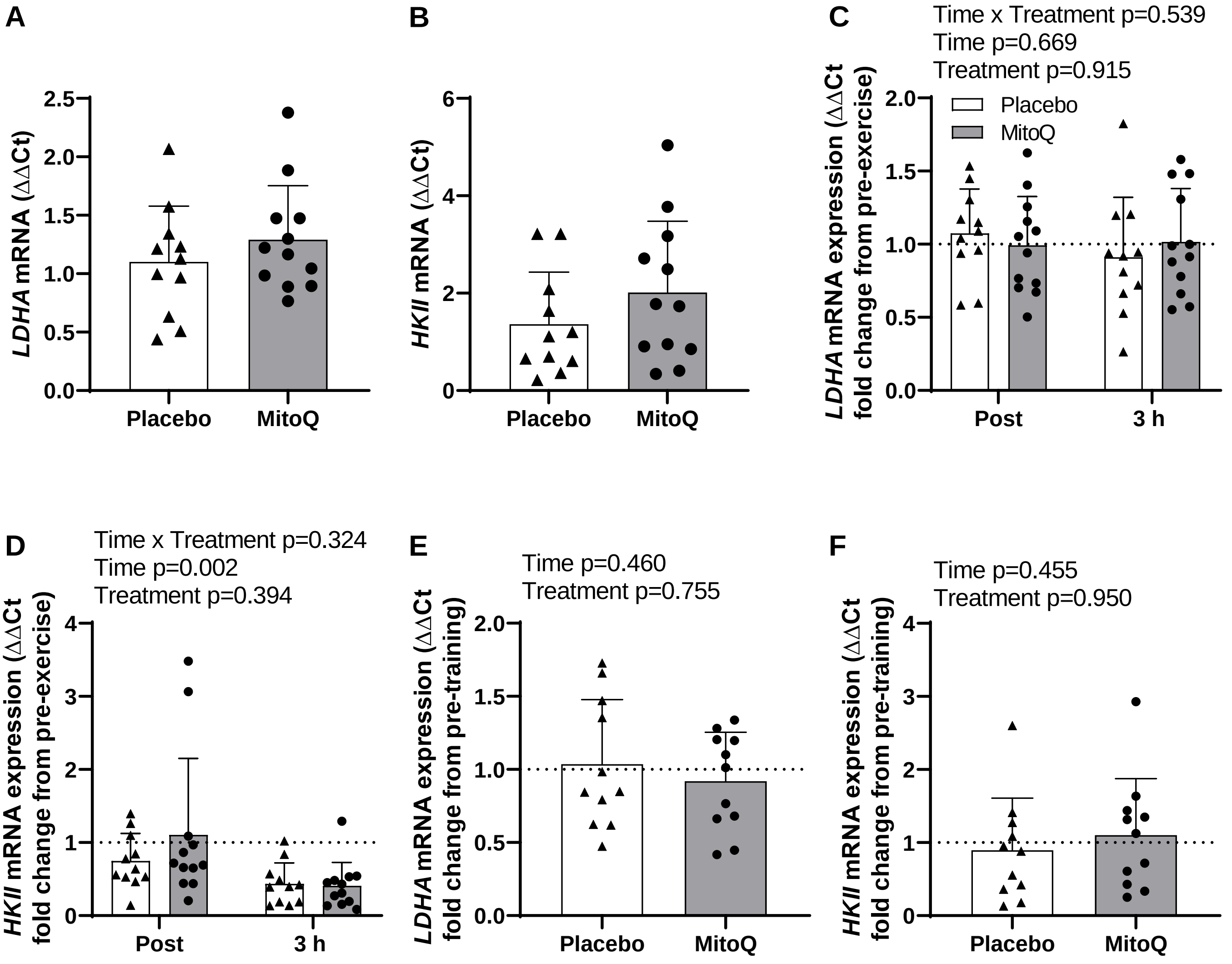
